# Geographical variation in treated psychotic and other mental disorders in Finland by region and urbanicity

**DOI:** 10.1101/2023.04.13.23288507

**Authors:** Kimmo Suokas, Olli Kurkela, Jaakko Nevalainen, Jaana Suvisaari, Christian Hakulinen, Olli Kampman, Sami Pirkola

**Author notes:** Corresponding author Kimmo Suokas, MD, Arvo Ylpön katu 34 (Arvo 1), 33014 University of Tampere, Finland.

## Abstract

**Background:** In Finland, prevalence of schizophrenia is higher in the eastern and northern regions and co-occurs with the distribution of schizophrenia polygenic risk scores. Both genetic and environmental factors have been hypothesized to contribute to this variation, but its reasons are not fully understood. We aimed to examine the prevalence of psychotic and other mental disorders by region and degree of urbanicity, and the impacts of socio-economic adjustments on these associations.

**Methods:** Nationwide population registers from 2011 to 2017 and healthcare registers from 1975 to 2017. We used 19 administrative and three aggregate regions, and a seven-level urban-rural classification with 250 × 250 m resolution. Prevalence ratios (PRs) were calculated by Poisson regression models and adjusted for gender, age, and calendar year (basic adjustments), and origin, residential history, urbanicity, household income, economic activity, and physical comorbidity (additional adjustments) on an individual level. Average marginal effects were used to visualize interaction effects between region and urbanicity.

**Results:** A total of 5 898 180 individuals were observed. All mental disorders were slightly more prevalent (PR 1.03 [95% CI, 1.02-1.03]), and psychotic disorders (1.11 [1.10-1.12]) and schizophrenia (1.19 [1.17-1.21]) considerably more prevalent in eastern and northern than in western coastal regions. After the additional adjustments, however, the PRs were 0.95 (0.95-0.96), 1.00 (0.99-1.01), and 1.03 (1.02-1.04), respectively. Urban residence was associated with increased prevalence of psychotic disorders across all regions (adjusted PR 1.21 [1.20-1.22]).

**Conclusions:** Socioeconomic and sociodemographic factors modulate the within-country geographical distribution of mental disorders highlighting their truly contextual nature.

## Background

The prevalence of psychotic and other mental disorders varies globally and locally (Simeone et al., 2015; GBD 2019 Mental Disorders Collaborators, 2022; Steel et al., 2014; Jongsma et al., 2018), with urban-rural differences being a particularly important factor in Northern Europe (Krabbendam et al., 2021; Pedersen et al., 2022; Vassos et al., 2016; Fett et al., 2019; Abrahamyan Empson et al., 2020). The underlying mechanisms for these variations are not well understood and are thought to be influenced by a combination of neighbourhood and individual-level social-environmental factors, including pollution, lack of green space, social stress or selective migration, among other things (Krabbendam et al., 2021). Some combined analyses have shown gene-environment synergism in the risk profiles (Colodro-Conde et al., 2018; Fan et al., 2018; Maxwell et al., 2021; Paksarian et al., 2018; Robinson & Bergen, 2021).

In Finland, there is a well-documented pattern of higher prevalence of schizophrenia and other psychotic disorders in the east and of mood and anxiety disorders in the south (Suvisaari et al., 2014; Perälä et al., 2008; Haukka et al., 2001; Korkeila et al., 1998; Hovatta et al., 1997; Lehtinen et al., 1990). In schizophrenia, regional differences have been more significant than urban-rural variations, and this geographical east-west pattern in schizophrenia prevalence coincides with schizophrenia polygenic risk scores, leading to the hypothesis that population genetics may play a role (see online Supplementary Fig. S1a)(Haukka et al., 2001; Kerminen et al., 2019; Kurki et al., 2019; Perälä et al., 2008). However, social determinants of mental health, such as the proportion of low-income earners (online Supplementary Fig. S1b), level of education, unemployment, migration, or household structure also vary across the country with less favourable compositions often seen in the eastern parts of the country. Urban areas, on the other hand, are more common in southern and western regions (online Supplementary Fig. S1c). It is not known to what extent regional and urban-rural variations interact, and to what extent the geographical variations are confounded by socioeconomic factors.

We aimed to evaluate regional and urban-rural variation in psychotic and all mental disorders, their interaction, and the impact of socioeconomic adjustments on these geographical differences. We hypothesized that much of the variability in prevalence of mental disorders would be explained by demographic and socioeconomic factors.

## Methods

We conducted a population-based register study including all individuals living in Finland from 2011 to 2017. Using individual-level population and health care registers, we calculated the prevalence of people with a history of mental health-related contact with primary care or psychiatric secondary inpatient or outpatient care on the last day of each of the study years. In addition, all individuals living in Finland between 1996 and 2017 were followed up in the registers to identify the incidence of the first psychiatric inpatient admissions. These time limits were based on the coverage of the national health care registers.

The Research Ethics Committee of the Finnish Institute for Health and Welfare approved the study protocol (decision #10/2016§751). Data were linked with permission from Statistics Finland (TK-53-1696-16) and the Finnish Institute of Health and Welfare. Informed consent is not required for register-based studies in Finland.

### Assessment of mental disorders

Information on mental healthcare was obtained from the Finnish Care Register for Health Care. Psychiatric inpatient care can be reliably recognized since 1975, secondary outpatient care has been included since 2008 and primary care has been included since 2011.

The International Statistical Classification of Diseases and Related Health Problems, Tenth Revision (ICD-10) has been used in Finland since 1996. We described specific disorders with the ten-level ICD-10 sub-chapter categories and in the following categories: all psychotic disorders (ICD-10: F20-29, F30.1, F30.2, F30.8, F30.9, F31.1, F31.2, F31.5, F31.6, F32.3, F33.3, F1x.5, F1x.7), mania and bipolar disorders with psychotic symptoms (F30.1, F30.2, F30.8, F30.9, F31.1, F31.2, F31.5, F31.6), psychotic depression (F32.3, F33.3), and substance-induced psychotic disorders (F1x.5, F1x.7). The diagnoses of schizophrenia and other primary psychotic disorders were classified in a particular order, with schizophrenia being the first (F20), followed by schizoaffective disorder (F25), delusional disorders (F22 and F24), brief psychotic disorders (F23), schizotypal disorder (F21), other nonorganic psychotic disorders (F28), and unspecified nonorganic psychosis (F29). If a person had more than one diagnosis from the schizophrenia spectrum, they were classified under the first group of disorders in the order presented above.

In primary care, *the ICPC-2 International Classification of Primary Care*, instead of ICD-10, is used in some facilities, and ICPC-2 mental health-related diagnoses were converted to corresponding ICD-10 sub-chapter categories when possible.

Discharge diagnoses and diagnoses from outpatient visits were also collected. A description of the method used for handling partly overlapping register data entries is publicly available (https://github.com/kmmsks/hilmo_identify_episodes/).

### Regions and urban-rural classification

Finland consists of 19 administrative regions, each with a central town, possible other towns and surrounding areas with varying degrees of urbanicity. Based on the distribution of the schizophrenia polygenic risk score (Kerminen et al., 2019; Kurki et al., 2019), we grouped the administrative regions into three aggregate regions: coastal, inland, and eastern and northern (online Supplementary Fig. S1d). The current region of residence on the last day of each study year was used for the main analysis. We used the seven-level urban-rural classification for the year 2010 issued by the Finnish Environment Institute (2013) based on a nationwide grid of 250 × 250 m cells, to measure urbanicity for each individual’s place of residence. In order to show geographical variation by region and urbanicity, we created maps with region-urbanicity subregions (online Supplementary Fig. S1c).

### Cofactors

We collected the following categorical individual-level demographic and socioeconomic data on the last day of each study year from the population registers: age (five-year intervals), gender (man or woman), origin (Finnish background or not), currently inhabiting the region of birth (yes or no), economic activity (employed; unemployed; students; pensioners and others outside the labour force), and equivalized household net income deciles. Net income was obtained after subtracting taxes and was adjusted for the size of the household dwelling unit using the Organisation for Economic Cooperation and Development–modified equivalence scale.

Physical comorbidity was assessed using the Charlson comorbidity index (CCI), a widely used comorbidity index with a weighted score of 17 comorbid conditions (Sundararajan et al., 2004). For each study year and for every individual in the study, the CCI score was calculated using available ICD-10 diagnoses of any actual treatment contact in healthcare registers from the beginning of the previous calendar year. Age was not included in the CCI scores but was adjusted in the main model. CCI scores were categorized by previously used cut-points: none, 1-3, and ≥4 (Erlangsen et al., 2020).

### Statistical analysis

The prevalence of a history of mental disorders was calculated for the last day of each calendar year of the study by summing the number of people with a history of mental health treatments in each region divided by the number of inhabitants in the region. Data were aggregated by strata defined by all possible combinations of cofactors. Prevalence ratios were examined using a Poisson regression model with a robust sandwich variance estimator. The strata in the aggregated data were taken as the unit of analysis and the log of population size of the strata was used as an offset term.

Regional prevalence ratios were adjusted for gender, age, and calendar year (basic adjustment). Additional adjustments for origin, residential history, urbanicity, household income, economic activity, and CCI were also made. Bayesian information criteria were used for the model selection.

For a fine-scale view of the variability of prevalence by region and urbanicity, the average marginal effects for each region-urbanicity subregion were predicted using a Poisson regression model that included a region-urbanicity interaction term. The predicted prevalence in each region-urbanicity subregion was calculated while holding the other predictors constant as observed (Williams, 2012).

The sensitivity to definitions of the outcome and explanatory variables was investigated by alternative definitions and comparison of results across the following additional analyses: The prevalence of all treated mental disorders and inpatient treatments only were compared; the incidence and prevalence of regional inpatient treatments were compared; and the current living region and the region of birth were compared. For data management and analyses, we used R, version 3.6.3 (R Project for Statistical Computing), and Stata, version 17.1 (StataCorp LLC).

## Results

During the years 2011 to 2017, a total of 5 898 180 individuals contributed to the study population. Altogether, 1 197 690 individuals of the total of 5 512 745 at the end of 2017 had a history of some medical contact in primary or secondary care mental health services. This resulted in a crude prevalence rate of 21.73% (24.07% in women and 19.32% in men). Prevalences stratified by the covariates are reported in the Supplementary Table.

The crude prevalence of all psychotic disorders, schizophrenia, and most of the other psychotic disorders was higher in the eastern and northern than in the coastal regions (Table). However, unspecified psychosis, bipolar disorder and substance-induced psychotic disorders, as well as mood disorders and neurotic disorders, were more common in the coastal region, resulting in only a minimal difference in the prevalence of all mental disorders (Table).

**Table.**
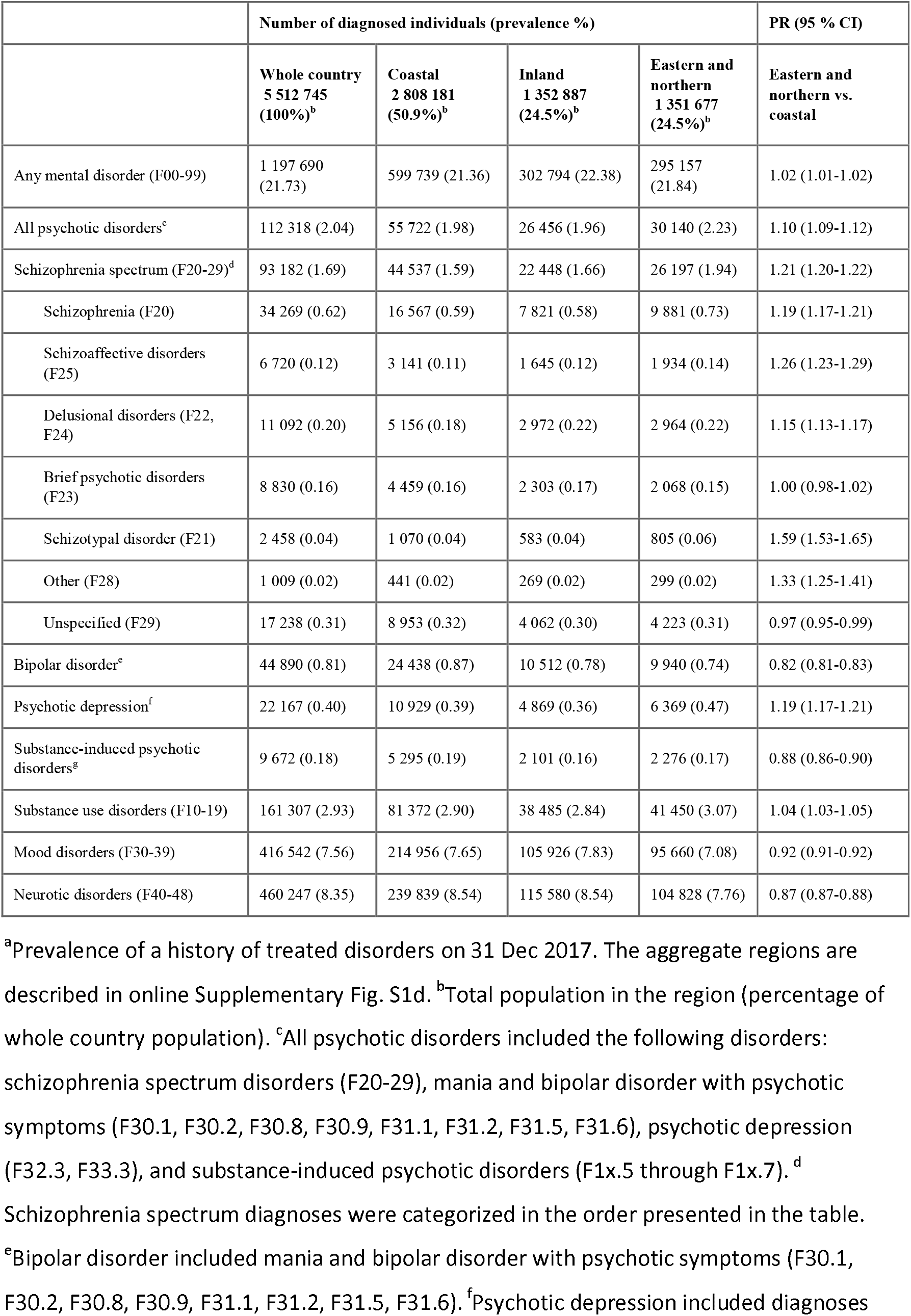
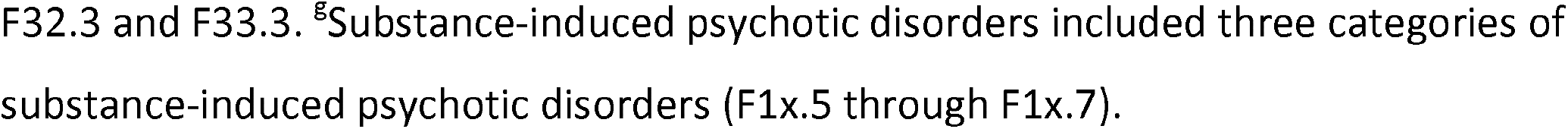
Prevalence of mental disorders by place of residence in 2017: number of cases, prevalence rates, and crude prevalence ratios (PR)^a^

|  | Number of diagnosed individuals (prevalence %) |  |  |  | PR (95 % CI) |
| --- | --- | --- | --- | --- | --- |
|  | Whole country<br>5 512 745<br>(100%)<sup>b</sup> | Coastal<br>2 808 181<br>(50.9%)<sup>b</sup> | Inland<br>1 352 887<br>(24.5%)<sup>b</sup> | Eastern and<br>northern<br>1 351 677<br>(24.5%)<sup>b</sup> | Eastern and<br>northern vs.<br>coastal |
| Any mental disorder (F00-99) | 1 197 690<br>(21.73) | 599 739 (21.36) | 302 794 (22.38) | 295 157<br>(21.84) | 1.02 (1.01-1.02) |
| All psychotic disorders<sup>c</sup> | 112 318 (2.04) | 55 722 (1.98) | 26 456 (1.96) | 30 140 (2.23) | 1.10 (1.09-1.12) |
| Schizophrenia spectrum (F20-29)<sup>d</sup> | 93 182 (1.69) | 44 537 (1.59) | 22 448 (1.66) | 26 197 (1.94) | 1.21 (1.20-1.22) |
| Schizophrenia (F20) | 34 269 (0.62) | 16 567 (0.59) | 7 821 (0.58) | 9 881 (0.73) | 1.19 (1.17-1.21) |
| Schizoaffective disorders (F25) | 6 720 (0.12) | 3 141 (0.11) | 1 645 (0.12) | 1 934 (0.14) | 1.26 (1.23-1.29) |
| Delusional disorders (F22, F24) | 11 092 (0.20) | 5 156 (0.18) | 2 972 (0.22) | 2 964 (0.22) | 1.15 (1.13-1.17) |
| Brief psychotic disorders (F23) | 8 830 (0.16) | 4 459 (0.16) | 2 303 (0.17) | 2 068 (0.15) | 1.00 (0.98-1.02) |
| Schizotypal disorder (F21) | 2 458 (0.04) | 1 070 (0.04) | 583 (0.04) | 805 (0.06) | 1.59 (1.53-1.65) |
| Other (F28) | 1 009 (0.02) | 441 (0.02) | 269 (0.02) | 299 (0.02) | 1.33 (1.25-1.41) |
| Unspecified (F29) | 17 238 (0.31) | 8 953 (0.32) | 4 062 (0.30) | 4 223 (0.31) | 0.97 (0.95-0.99) |
| Bipolar disorder<sup>e</sup> | 44 890 (0.81) | 24 438 (0.87) | 10 512 (0.78) | 9 940 (0.74) | 0.82 (0.81-0.83) |
| Psychotic depression<sup>f</sup> | 22 167 (0.40) | 10 929 (0.39) | 4 869 (0.36) | 6 369 (0.47) | 1.19 (1.17-1.21) |
| Substance-induced psychotic disorders<sup>g</sup> | 9 672 (0.18) | 5 295 (0.19) | 2 101 (0.16) | 2 276 (0.17) | 0.88 (0.86-0.90) |
| Substance use disorders (F10-19) | 161 307 (2.93) | 81 372 (2.90) | 38 485 (2.84) | 41 450 (3.07) | 1.04 (1.03-1.05) |
| Mood disorders (F30-39) | 416 542 (7.56) | 214 956 (7.65) | 105 926 (7.83) | 95 660 (7.08) | 0.92 (0.91-0.92) |
| Neurotic disorders (F40-48) | 460 247 (8.35) | 239 839 (8.54) | 115 580 (8.54) | 104 828 (7.76) | 0.87 (0.87-0.88) |

<sup>a</sup>Prevalence of a history of treated disorders on 31 Dec 2017. The aggregate regions are described in online Supplementary Fig. S1d. <sup>b</sup>Total population in the region (percentage of whole country population). <sup>c</sup>All psychotic disorders included the following disorders: schizophrenia spectrum disorders (F20-29), mania and bipolar disorder with psychotic symptoms (F30.1, F30.2, F30.8, F30.9, F31.1, F31.2, F31.5, F31.6), psychotic depression (F32.3, F33.3), and substance-induced psychotic disorders (F1x.5 through F1x.7). <sup>d</sup>Schizophrenia spectrum diagnoses were categorized in the order presented in the table. <sup>e</sup>Bipolar disorder included mania and bipolar disorder with psychotic symptoms (F30.1, F30.2, F30.8, F30.9, F31.1, F31.2, F31.5, F31.6). <sup>f</sup>Psychotic depression included diagnoses
F32.3 and F33.3. <sup>8</sup>Substance-induced psychotic disorders included three categories of substance-induced psychotic disorders (F1x.5 through F1x.7).

After basic adjustments, prevalence ratios (PRs) of 1.11 (95% CI, 1.10-1.12) for all psychotic disorders, 1.20 (1.19-1.21) for schizophrenia spectrum, 0.85 (0.84-0.86) for bipolar disorder, and 1.03 (1.02-1.03) for all mental disorders in the eastern and northern compared to the coastal regions were observed (Fig. 1a).

**Fig. 1.**
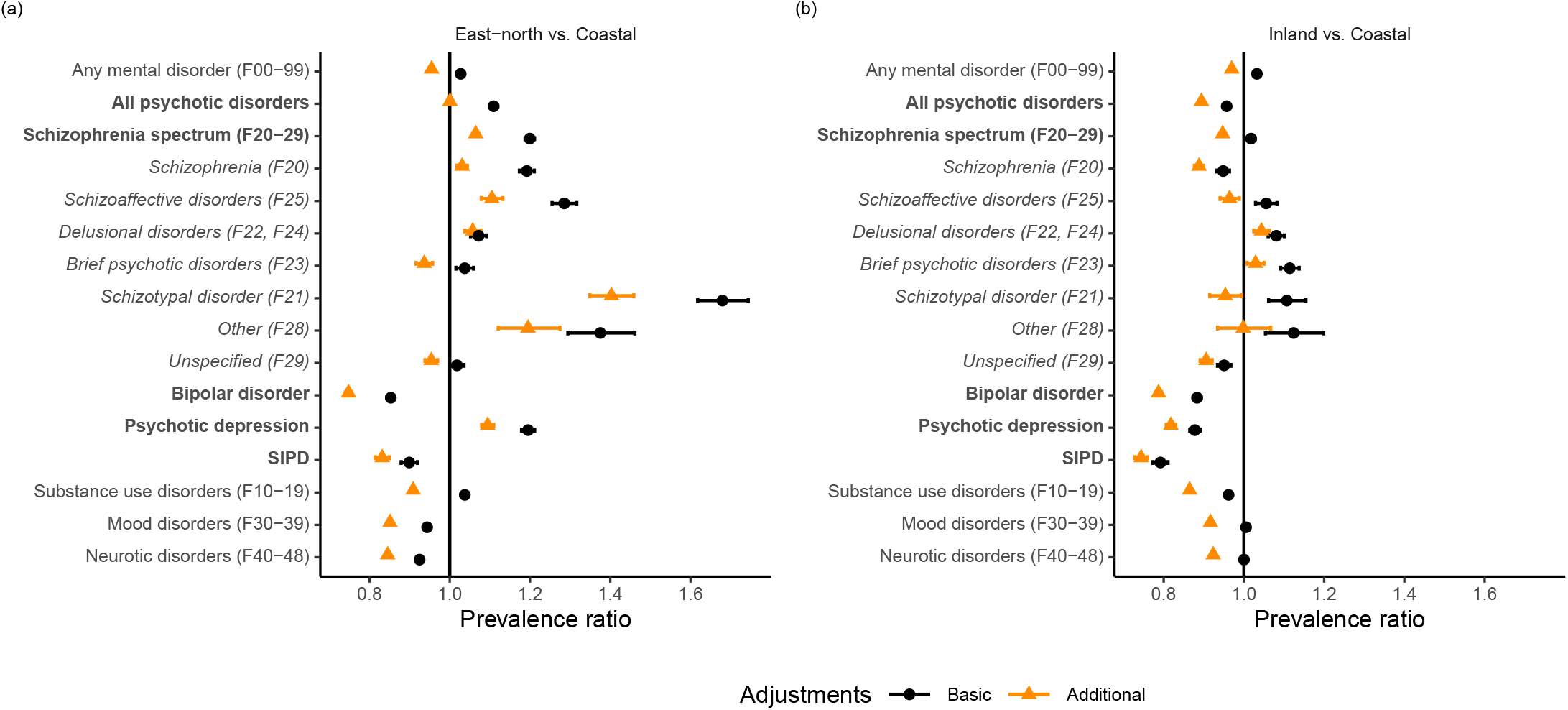
Prevalence ratios of mental disorders by place of residence. (a) eastern and northern vs. coastal and (b) inland vs. coastal. Coastal, Inland, and East-north regions are described in online Supplementary Fig. S1d. In the basic adjustment, prevalence ratios are adjusted for age, gender, and calendar time. In the additional adjustment, prevalence ratios were adjusted for age, gender, calendar time, urbanicity, origin, residence history, household income, economic activity, and Charlson comorbidity index. Error bars indicate 95% CIs. Subgroups of all included psychotic disorders are highlighted in bold. Bipolar disorder included ICD-10 codes F30.1, F30.2, F30.8, F30.9, F31.1, F31.2, F31.5, F31.6, psychotic depression codes F32.3 and F33.3, and substance-induced psychotic disorders codes F1x.5 to F1x.7. Schizophrenia spectrum diagnoses (in italic) were categorized in the order presented in the figure.

When additional adjustments for socioeconomic factors and comorbidities were included in the models, the eastern and northern prominence in psychotic disorders disappeared, with a PR of 1.00 (0.99-1.01). PRs of 1.06 (1.06-1.07) for schizophrenia spectrum, 1.03 (1.02-1.04) for schizophrenia, and 0.75 (0.74-0.76) for bipolar disorder were observed (Fig. 1a). The PR for all mental disorders was 0.95 (0.95-0.96) (Fig. 1a). Adding income to the models caused a major change in the PR estimates, and the effect of each of the additional covariates is shown in the online Supplementary Fig S2. There were some variations between neighbouring regions within the aggregate regions and between diagnoses (online Supplementary Fig. S3).

Residence in inner urban areas or in the local centres in rural areas was clearly associated with increased prevalence of all mental disorders and major psychotic disorders in both levels of adjustment (Fig. 2). The additional adjustments changed the prevalence ratios in some levels of urbanicity, although the link between urbanicity and psychotic disorders remained clear. In inner urban areas, PRs of 1.10 (1.10-1.10) for all mental disorders and 1.21 (95% CI, 1.20-1.22) for psychoses, compared to the whole national mean with additional adjustments, were observed.

**Fig. 2.**
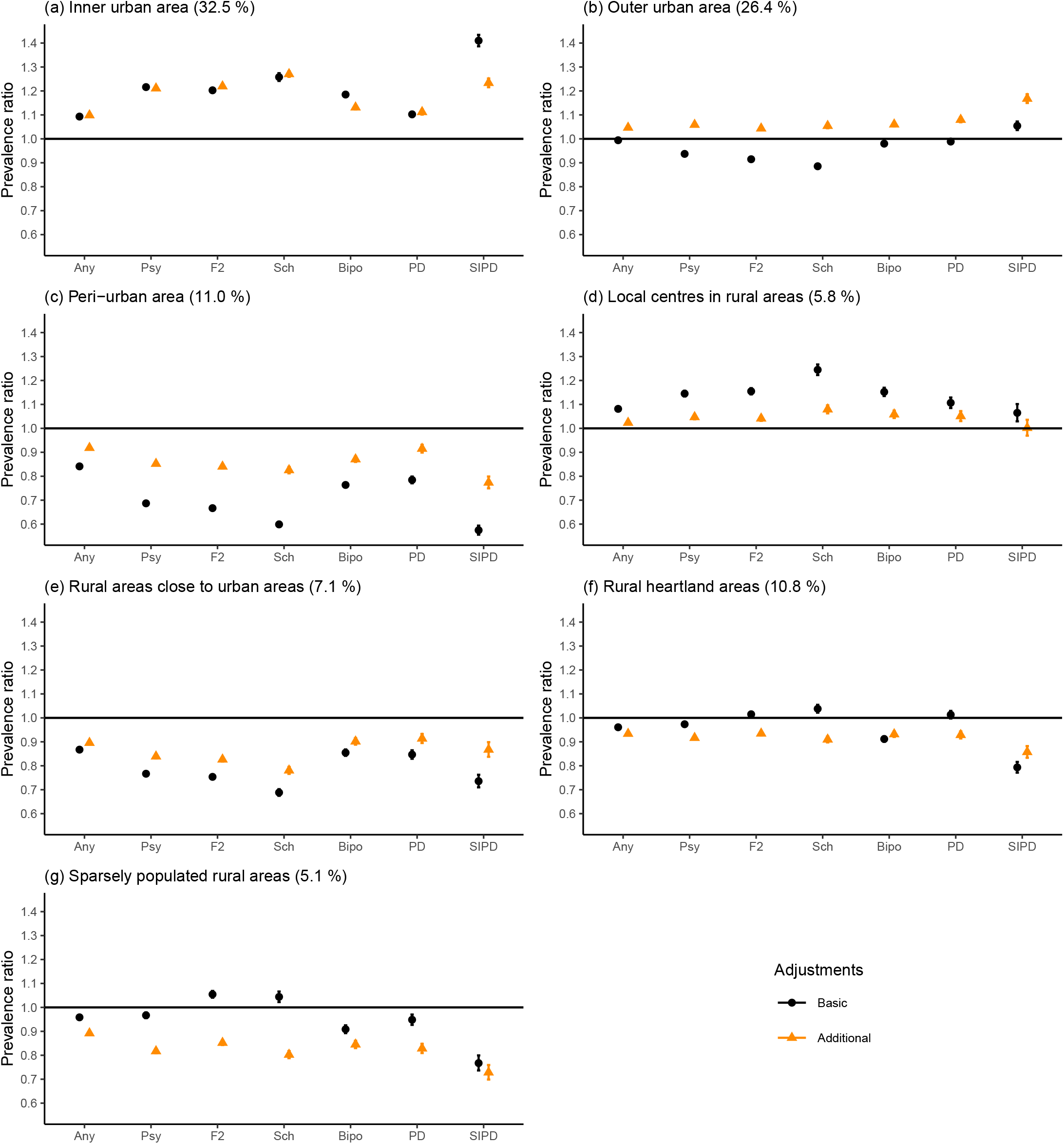
Prevalence ratios of selected mental disorders by urbanicity of the place of residence, compared to the national mean. The proportion of population living in each level of urbanicity is given in parentheses. Any refers to any mental disorder, Psych to all psychotic disorders, F2 to schizophrenia spectrum, Sch to schizophrenia, Bipo to bipolar disorder, PDe to psychotic depression, and SIPD refers to substance-induced psychotic disorders. In the basic adjustment, prevalence ratios are adjusted for age, gender, and calendar time. In the additional adjustment, prevalence ratios were adjusted for age, gender, calendar time, region, origin, residence history, household income, economic activity, and Charlson comorbidity index. Error bars indicate 95% CIs.

The analysis of prevalence of mental disorders by region of residence and urbanicity with basic adjustments showed an eastern and northern prominence in the prevalence of all mental disorders and psychotic disorders in all levels of urbanicity. After the additional adjustments, prominence of the urban areas in the coastal regions became evident (online Supplementary Fig. S4). The average marginal effects of prevalence for each region-urbanicity subregion are visualized in the maps (Fig. 3 and Fig. 4).

**Fig. 3.**
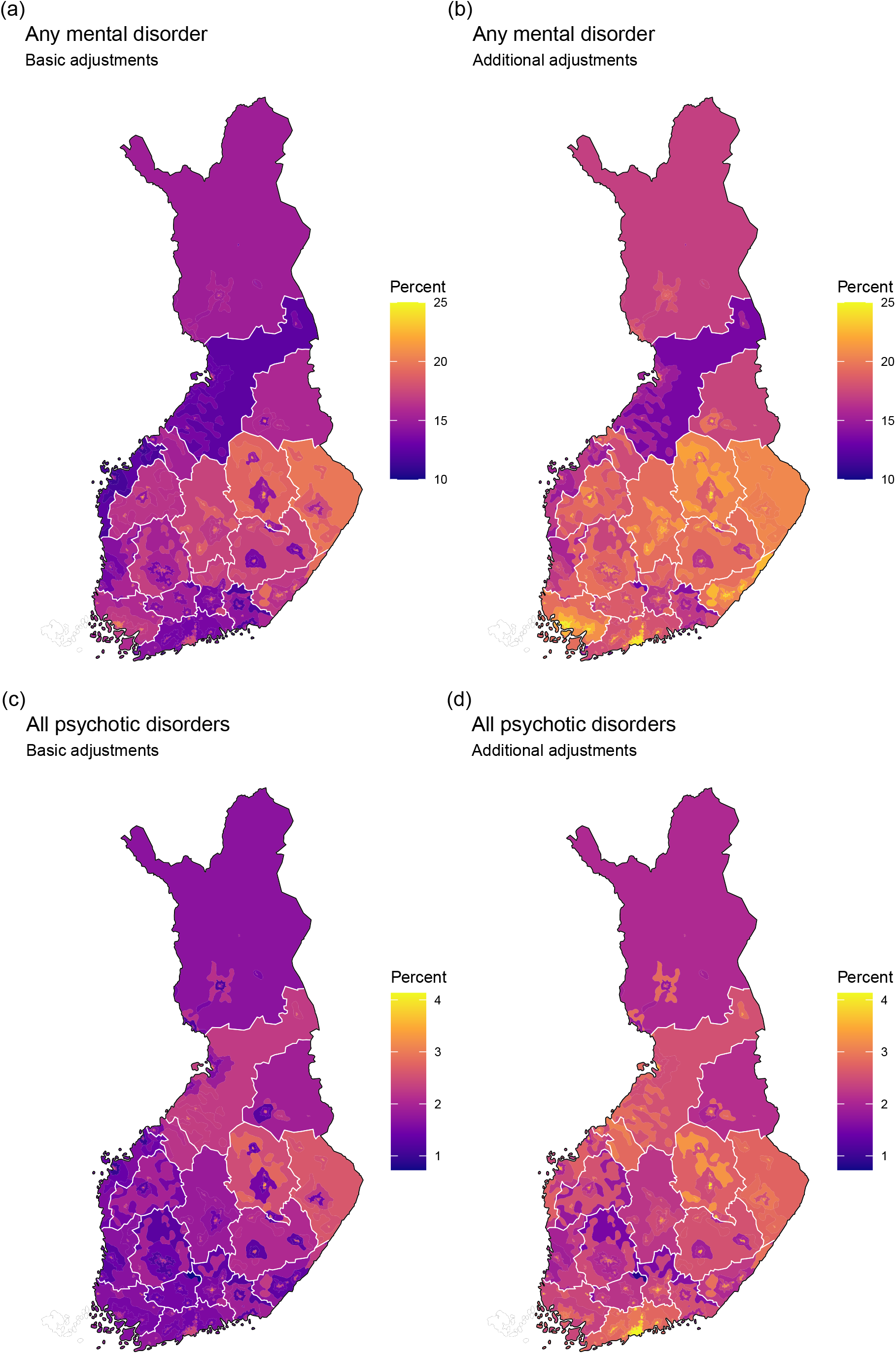
Average marginal effects of region of residence and urbanicity on the prevalence of any mental disorder and all psychotic disorders. (a) any mental disorder, basic adjustments, (b) any mental disorder, additional adjustments, (c) all psychotic disorders, basic adjustments, and (d) all psychotic disorders, additional adjustments. Predicted prevalence in each region-urbanicity subregion was calculated while holding the other predictors constant as observed. In the basic adjustment, prevalence ratios were adjusted for age, gender, and calendar time. In the additional adjustment, prevalence ratios were adjusted for age, gender, calendar time, origin, residence history, household income, economic activity, and Charlson comorbidity index.

**Fig. 4:**
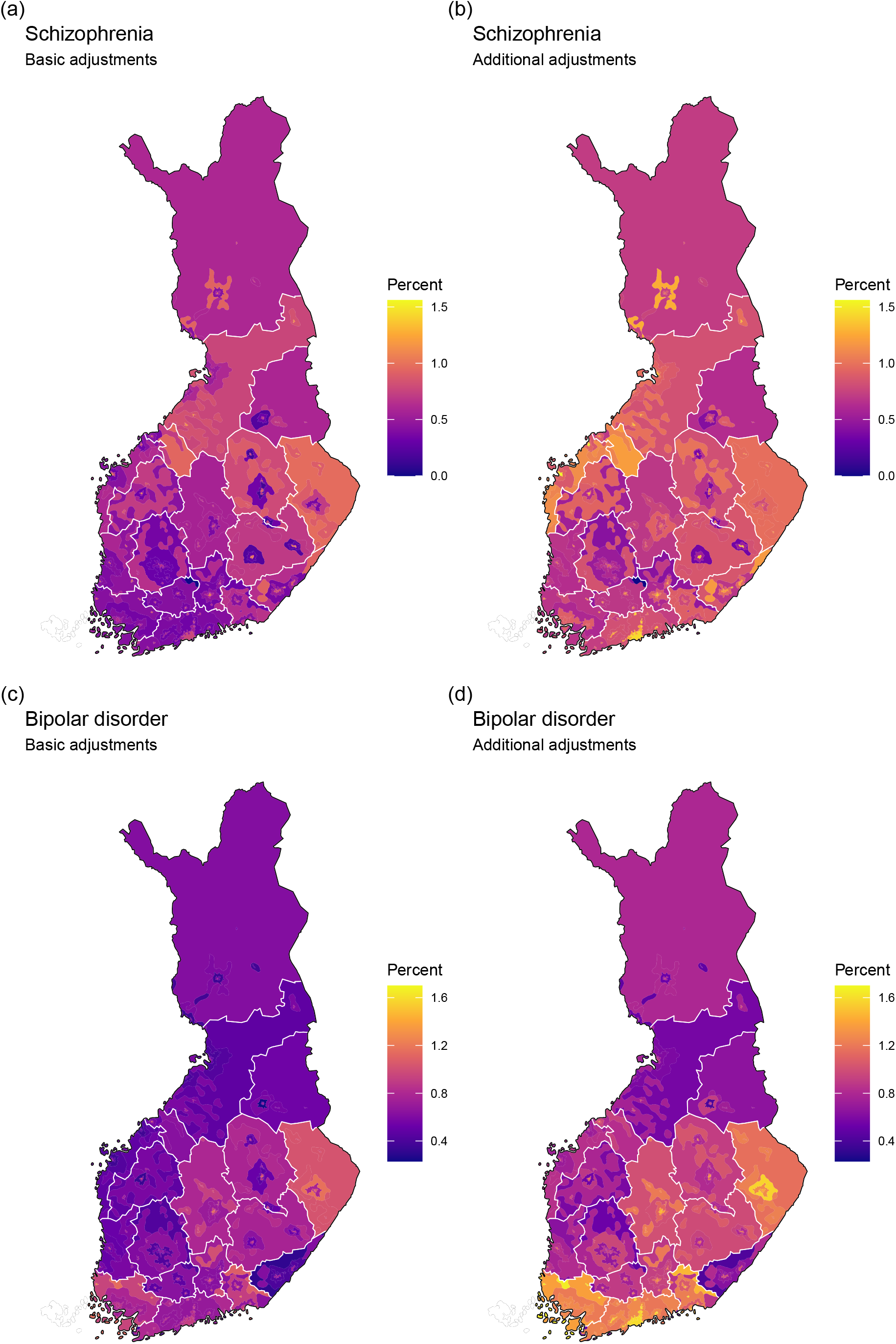
Average marginal effects of region of residence and urbanicity on the prevalence of schizophrenia and bipolar disorder. (a) schizophrenia, basic adjustments, (b) schizophrenia, additional adjustments, (c) bipolar disorder, basic adjustments, and (d) bipolar disorder, additional adjustments. Predicted prevalence in each region-urbanicity subregion was calculated while holding the other predictors constant as observed. In the basic adjustment, prevalence ratios were adjusted for age, gender, and calendar time. In the additional adjustment, prevalence ratios were adjusted for age, gender, calendar time, origin, residence history, household income, economic activity, and Charlson comorbidity index.

The following additional analyses were conducted: First, if inpatient care was analyzed alone, clear eastern and northern prominence would have been observed (online Supplementary Fig. S5). Second, using region of birth instead of the region of residence would not alter the overall findings (online Supplementary Fig. S6). Third, using data on incidence of the first inpatient episodes instead of prevalence would cause minor changes in the proportions of different diagnostic categories, but would not change the overall findings (online Supplementary Fig. S7). Fourth, if men and women were analyzed separately, the main finding would not change (online Supplementary Fig. S8).

## Discussion

In this nationwide register-based study of over 5 million Finnish persons, we found that the prevalence of all mental disorders and psychotic disorders treated in both primary or secondary care was higher in the eastern and northern regions compared to coastal regions. After adjusting for socioeconomic factors, however, this geographical difference was no longer evident. By contrast, the urban-rural differences, as measured using a detailed seven-level classification of current residency, persisted after the adjustments and were consistent with previous findings from other Nordic countries. Urban effect was evident across the country and diagnostic categories, although regional differences in some diagnostic subgroups, such as schizophrenia and bipolar disorders, were observed. Taken together, our results demonstrate the significant impact of social determinants on the mental health of the population and have important national implications.

To the best of our knowledge, this is the first comprehensive study demonstrating the associations between the within-country distribution of socioeconomic and demographic factors and the prevalence of mental disorders treated with either primary or secondary care. Epidemiological studies in Finland have investigated regional and urban-rural variations in mental disorders since the 1930s (Haukka et al., 2001; Hovatta et al., 1997; Korkeila et al., 1998; Lehtinen et al., 1990; *Mielisairaat ja vajaamieliset*, 1940; Perälä et al., 2008; Suvisaari et al., 2014). The east-west differences have been consistently observed, but one recent study found significant regional variation in mental disorder disability pensions that did not follow the traditional east-west health differences (Karolaakso et al., 2021). Thus, within-country geographical differences in mental health are sensitive to a variety of social determinants and draw a more complex picture than reported in previous studies.

Previous findings on the association between urbanicity and mental disorders in Finland have been mixed, with the earliest studies showing an association between living in cities and schizophrenia (*Mielisairaat ja vajaamieliset*, 1940), but more recent studies suggestive of urban effects but yielding inconsistent results (Haukka et al., 2001; Perälä et al., 2008; Suvisaari et al., 2014). Current results align with previous studies in Northern Europe, demonstrating an association between variety of psychotic disorders and urbanicity (Krabbendam et al., 2021; Plana-Ripoll et al., 2018; Vassos et al., 2016). Finland no longer appears to be an exception in this respect. Structural changes in demography, employment and services have affected particularly eastern rural parts of the country in recent decades and probably affect the temporal differences in the link between urbanicity and psychotic disorders in Finland (Haukka et al., 2001; Makkonen et al., 2022).

Household income was a particularly strong cofactor in the models. This is not surprising, as income inequality and individual level low income and mental disorders have been strongly linked with complex bi-directional pathways (Pickett & Wilkinson, 2015; Hakulinen et al., 2020; Ridley et al., 2020; Suokas et al., 2020; Hakulinen et al., 2023). In the current study, we did not explore the causal pathways behind the mental disorders and income distribution. Nevertheless, income was a relevant cofactor, as it is unlikely that the within-country distribution of income was determined by the regional prevalence of mental disorders.

Contrary to regional differences, urban-rural variation did not disappear after socioeconomic adjustments. Urban environments in a sparsely populated country such as Finland may vary greatly within the country in terms of potential urban risk attributes such as nature spaces, migration, social stress, or demographical and socioeconomic composition. Our analysis of the urbanicity-region interaction with socioeconomic adjustments showed that urbanicity is a relevant factor for mental health in all regions of the country, regardless of the size of the regional urban centre, from Kajaani with a population of 36,000 to Helsinki with a population of 665,000. We evaluated regional differences and urbanicity based on current residency, while controlling for living in the birth region. This approach enabled accounting for within-country migration. However, we did not have data on individual histories of urban residency or changes in geographical distribution of urbanicity. Selective migration can affect regional composition and socioeconomic contexts, and also affects the associations between urbanicity and psychotic disorders (Colodro-Conde et al., 2018; Krabbendam et al., 2021; Pedersen, 2015; Sariaslan et al., 2016). In Finland, however, it has been suggested that individuals with mental disorders are not particularly likely to move to the most urban centers (Vaalavuo & Sihvola, 2021).

The relatively high prevalence of bipolar disorder with psychotic features in southern urban areas and the comparatively high prevalence of schizophrenia spectrum diagnoses in eastern and northern areas emphasize the importance of considering different register-based diagnoses side by side. Although the Finnish registers show good consistence (Sund, 2012), register-based studies have shown higher incidence rates of psychotic disorders than first-contact designs, which may relate partly to diagnostic practices (Jongsma et al., 2019), and a tendency towards a narrow definition of schizophrenia in clinical practice in Finland has been recognized (Isohanni et al., 1997). Whether there are differences in diagnostic practices in primary or secondary care mental health services across the country has not been evaluated recently. With the observed differences in certain diagnostic categories in mind, assessing real-world diagnostic consistency and reliability might be useful in terms of both scientific and clinical accuracy.

The study of population genetics in Finland has attracted a great deal of interest, and there is a well-documented north-south and east-west genetic differentiation within the population (Kerminen et al., 2017; Kurki et al., 2019, 2023). Although the use of polygenic risk scores for explaining geographic differences in phenotypes is not currently recommended due to methodological limitations, the striking similarity between schizophrenia prevalence and polygenic scores has been suggested as an example of the potential of polygenic risk scores to explain geographic health differences (Kerminen et al., 2019). Our results showed that after adjusting for socioeconomic factors, the prevalence of all psychotic disorders did not display statistically significant east-west differences, and did not align with the geographical gradient of schizophrenia polygenic scores. A diagnosis of schizophrenia was slightly more prevalent in eastern parts of the country, but did not follow a gradient that was comparable to that of schizophrenia polygenic scores. Thus, accounting for neighbourhood contextual factors and socioeconomic composition and individual level social determinants, together with genetic information, may be beneficial in future studies of geographical differences in mental health in Finland.

### Strengths and limitations

The main strength of our study is the use of interlinked Finnish national registers, which provide comprehensive data on both primary and secondary care treatments for mental disorders across the country. The inclusion of primary care treatment data is important, as primary care mental health treatment is common in Finland, and our previous study showed that including primary care may alter findings (Suokas et al., 2022). There is no universal definition of urbanicity, and to the best of our knowledge the current seven-level classification with 250 × 250 m pixels has not been used before in this context and is more detailed than previous classifications.

This study has certain limitations. First, primary care data is available only since 2011. Hence, incident cases cannot be recognized. The prevalence of mental health treatments was the outcome of interest, and we did not focus on the complex bi-directional causal chains of income and mental health on an individual level, but rather on the overall composition of the population. Second, the current urban-rural classification is available only since 2010, and therefore historical changes in urban effects cannot be evaluated and the individual level residence history by urbanicity cannot by traced. Third, no individual level genetic data was used and thus the comparison between our study and that of Kurki et al. (2019) is indirect. Fourth, private and employer-paid mental health outpatient care are significant components of the Finnish health care system, and probably more common in urban settings, but were not covered in the registers for the study period. Finally, the present observational results do not have causal interpretation.

## Conclusion

Urbanicity and socioeconomic position are important determinants of geographical variations in population mental health. In this study, the previously well documented east-west gradient in psychotic disorders that coincides with the geographical distribution of schizophrenia polygenic risk scores, was no longer observed after detailed adjustments. Our current findings align with previous studies in Northern Europe, demonstrating a solid association between psychotic disorders and urbanicity also in Finland, which has previously been uncertain. Further study is needed to provide better understanding of the geographical patterns of mental health.

## Data Availability

The data that support the findings of this study are available from the National Institute of Health and Welfare (http://www.thl.fi) and Statistics Finland (www.stat.fi). Restrictions apply to the availability of these data, which were used under license for this study. Inquiries about secure access to data should be directed to data permit authority Findata (https://findata.fi/en/).

## Funding Statement

This study was funded by the Jalmari and Rauha Ahokas Foundation, European Research Council (ERC) under the European Union’s Horizon 2020 research and innovation programme (Grant agreement No. 101040247 to CH), and the Finnish Psychiatric Association. The funding sources had no role in study design, data collection, analysis, interpretation, writing, or submission.

## Conflict of interest

We declare no competing interests.

## Notes

**Financial support:** Jalmari and Rauha Ahokas Foundation, European Research Council (ERC) under the European Union’s Horizon 2020 research and innovation programme (Grant agreement No. 101040247 to CH), and the Finnish Psychiatric Association.

### Competing Interest Statement

The authors have declared no competing interest.

### Author Declarations

Ethics Committee of the Finnish Institute for Health and Welfare gave ethical approval for this work.

