## Supplementary data for "Geographical variation in treated psychotic and other mental disorders in Finland by region and urbanicity"

### Supplementary material

**Supplementary Figure S1: Regional compositions of Finland**

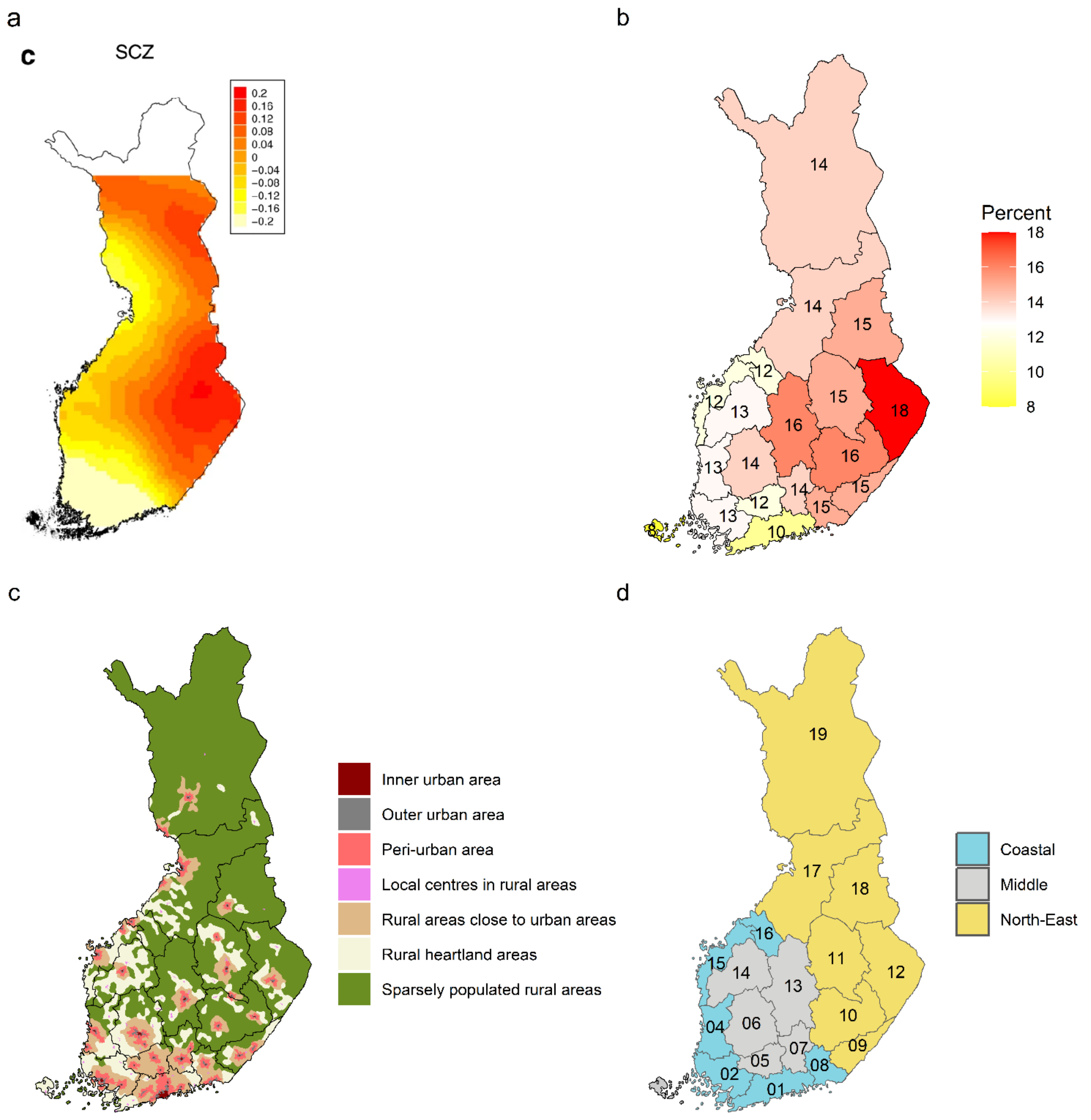

a Regional distribution of schizophrenia polygenic risk-score. Source: Figure 6c in Kurki et al. (2019). Available under a Creative Commons Attribution 4.0 License.

b Low-income earners (percentage of persons at risk of poverty) by region. Source: Statistics Finland (2017)

c Urban-rural classification and administrative regions in Finland. Source: Finnish Environment Institute (2013)

d Administrative regions in Finland and aggregate regions based on the polygenic risk-score distribution in this study. Regions: 01 Uusimaa, 02 Varsinais-Suomi, 04 Satakunta, 05 Kanta-Häme, 06 Pirkanmaa, 07 Päijät-Häme, 08 Kymenlaakso, 09 South Karelia, 10 Etelä-Savo, 11 Pohjois-Savo, 12 North Karelia, 13 Central Finland, 14 South Ostrobothnia, 15 Ostrobothnia, 16 Central Ostrobothnia, 17 North Ostrobothnia, 18 Kainuu, 19 Lapland

**Supplementary Table: Prevalence of all mental disorders stratified by the cofactors**

| | **Whole country** | | |
|---|---|---|---|
| | **N (%)** | **Population (%)** | **Prevalence, %** |
| Gender | | | |
| Men | 525 338 (43.9) | 2 719 032 (49.3) | 19.3 |
| Women | 672 352 (56.1) | 2 793 713 (50.7) | 24.1 |
| Person's origin | | | |
| Finnish background, born in Finland | 1 120 105 (93.5) | 5 077 539 (92.1) | 22.1 |
| Finnish background, born abroad | 14 604 (1.2) | 51 385 (0.9) | 28.4 |
| Foreign background, born in Finland | 11 377 (0.9) | 62 572 (1.1) | 18.2 |
| Foreign background, born abroad | 51 604 (4.3) | 321 249 (5.8) | 16.1 |
| Living in the region of birth | | | |
| No | 438 742 (36.6) | 1 948 735 (35.3) | 22.5 |
| Yes | 758 948 (63.4) | 3 564 010 (64.7) | 21.3 |
| Urbanicity | | | |
| Inner urban area | 428 063 (35.7) | 1 792 079 (32.5) | 23.9 |
| Outer urban area | 308 122 (25.7) | 1 454 480 (26.4) | 21.2 |
| Peri-urban area | 109 262 (9.1) | 608 651 (11.0) | 18.0 |
| Local centres in rural areas | 73 324 (6.1) | 318 981 (5.8) | 23.0 |
| Rural areas close to urban areas | 71 120 (5.9) | 388 748 (7.1) | 18.3 |
| Rural heartland areas | 120 886 (10.1) | 593 821 (10.8) | 20.4 |
| Sparsely populated rural areas | 57 795 (4.8) | 283 320 (5.1) | 20.4 |
| Missing | 29 118 (2.4) | 72 665 (1.3) | 40.1 |
| Household income decile | | | |
| Non-dwelling | 54 208 (4.5) | 191 679 (3.5) | 28.3 |
| 1 (lowest) | 202 106 (16.9) | 508 435 (9.2) | 39.8 |
| 2 | 161 197 (13.5) | 525 510 (9.5) | 30.7 |
| 3 | 130 224 (10.9) | 526 325 (9.5) | 24.7 |
| 4 | 113 879 (9.5) | 526 957 (9.6) | 21.6 |
| 5 | 105 669 (8.8) | 527 813 (9.6) | 20.0 |
| 6 | 99 042 (8.3) | 528 182 (9.6) | 18.8 |
| 7 | 91 648 (7.7) | 528 805 (9.6) | 17.3 |
| 8 | 84 885 (7.1) | 529 895 (9.6) | 16.0 |

| | Whole country | | |
|---|---|---|---|
| | **N (%)** | **Population (%)** | **Prevalence, %** |
| 9 | 75 044 (6.3) | 531 314 (9.6) | 14.1 |
| 10 (highest) | 58 336 (4.9) | 532 233 (9.7) | 11.0 |
| Data missing | 21 452 (1.8) | 55 597 (1.0) | 38.6 |
| Economic activity | | | |
| Employed | 411 940 (34.4) | 2 327 641 (42.2) | 17.7 |
| Unemployed | 113 970 (9.5) | 296 457 (5.4) | 38.4 |
| Aged 0 to 14 | 133 651 (11.2) | 890 376 (16.2) | 15.0 |
| Student, pupil | 115 233 (9.6) | 400 761 (7.3) | 28.8 |
| Pensioners and others outside the labour force | 422 896 (35.3) | 1 597 510 (29.0) | 26.5 |
| Charlson comorbidity index | | | |
| None | 1 003 981 (83.8) | 4 880 244 (88.5) | 20.6 |
| 1-3 | 182 543 (15.2) | 596 759 (10.8) | 30.6 |
| ≥4 | 11 166 (0.9) | 35 742 (0.6) | 31.2 |

**Supplementary Figure S2: Prevalence ratios of selected mental disorders by place of residence with different levels of adjustment**

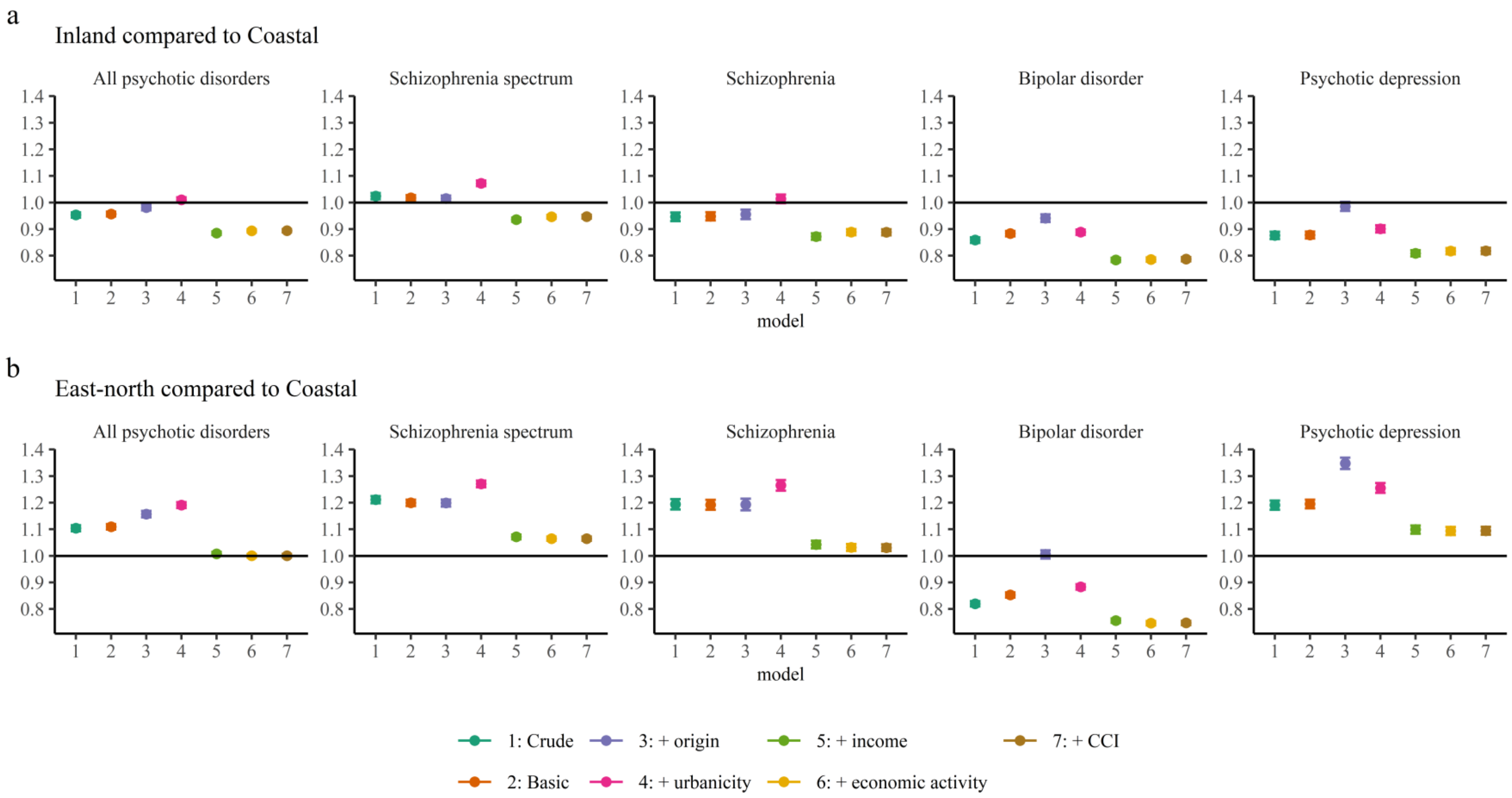

Coastal and eastern and northern regions are described in Figure 1d. In the basic adjustment, prevalence ratios are adjusted for age, gender, and calendar time. Origin refers to persons living in their region of birth. CCI refers to Charlson comorbidity index. Error bars indicate 95% Cis.

**Supplementary Figure S3: Prevalence ratios of selected mental disorders by place of residence in different administrative regions of Finland, compared to the national mean**

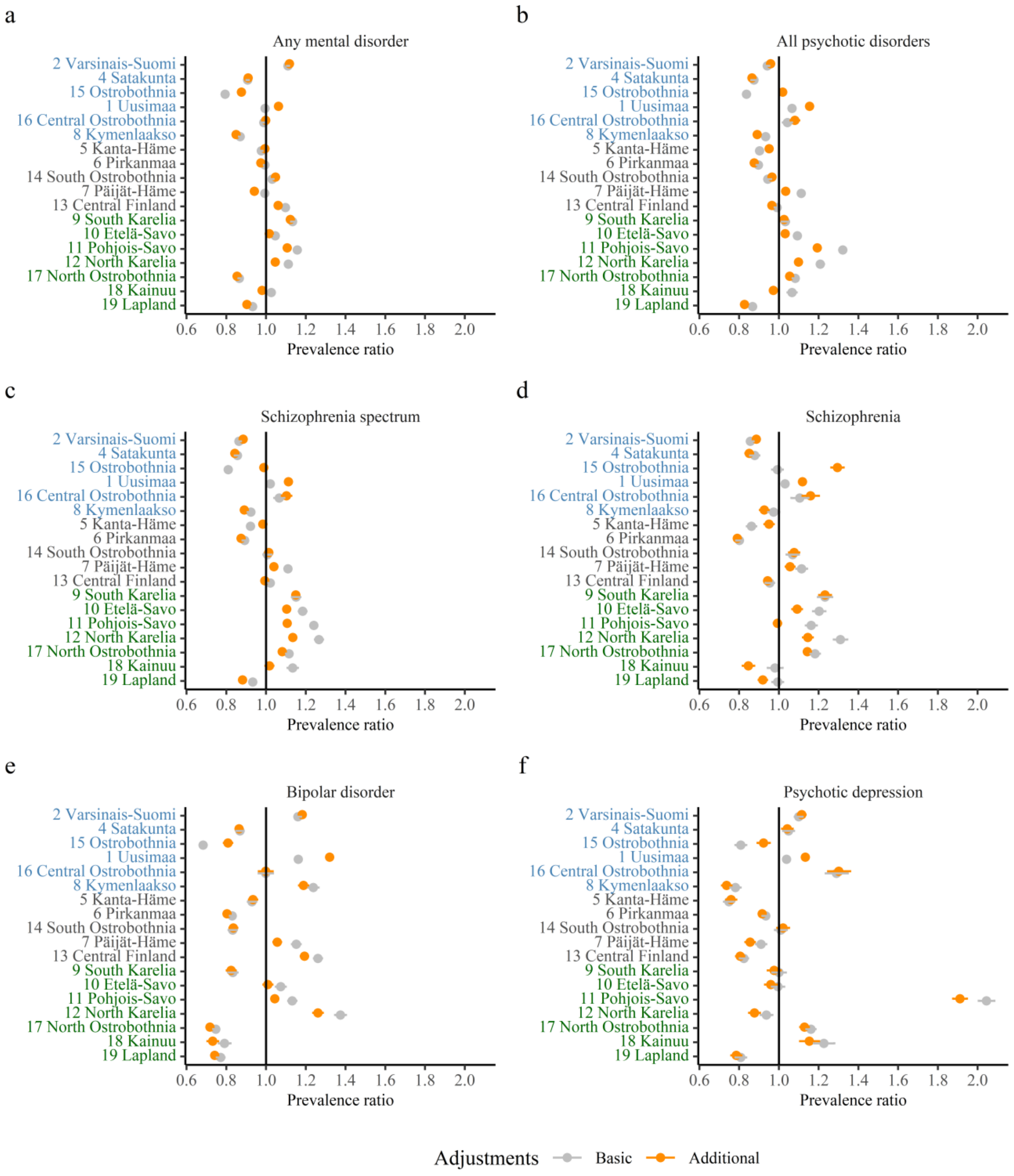

Numbers of the regions correspond to the numbers in the map in Figure 1d. Coastal regions are highlighted in blue, eastern and northern parts in green. In the basic adjustment, prevalence ratios are adjusted for age, gender, and calendar time. In the additional adjustment, prevalence ratios are adjusted for age, gender, calendar time, urbanicity, origin, residence history, household income, economic activity, and Charlson comorbidity index. Error bars indicate 95% CIs.

**Supplementary Figure S4: Average marginal effects of aggregated region of residence and urbanicity on the prevalence of mental disorders**

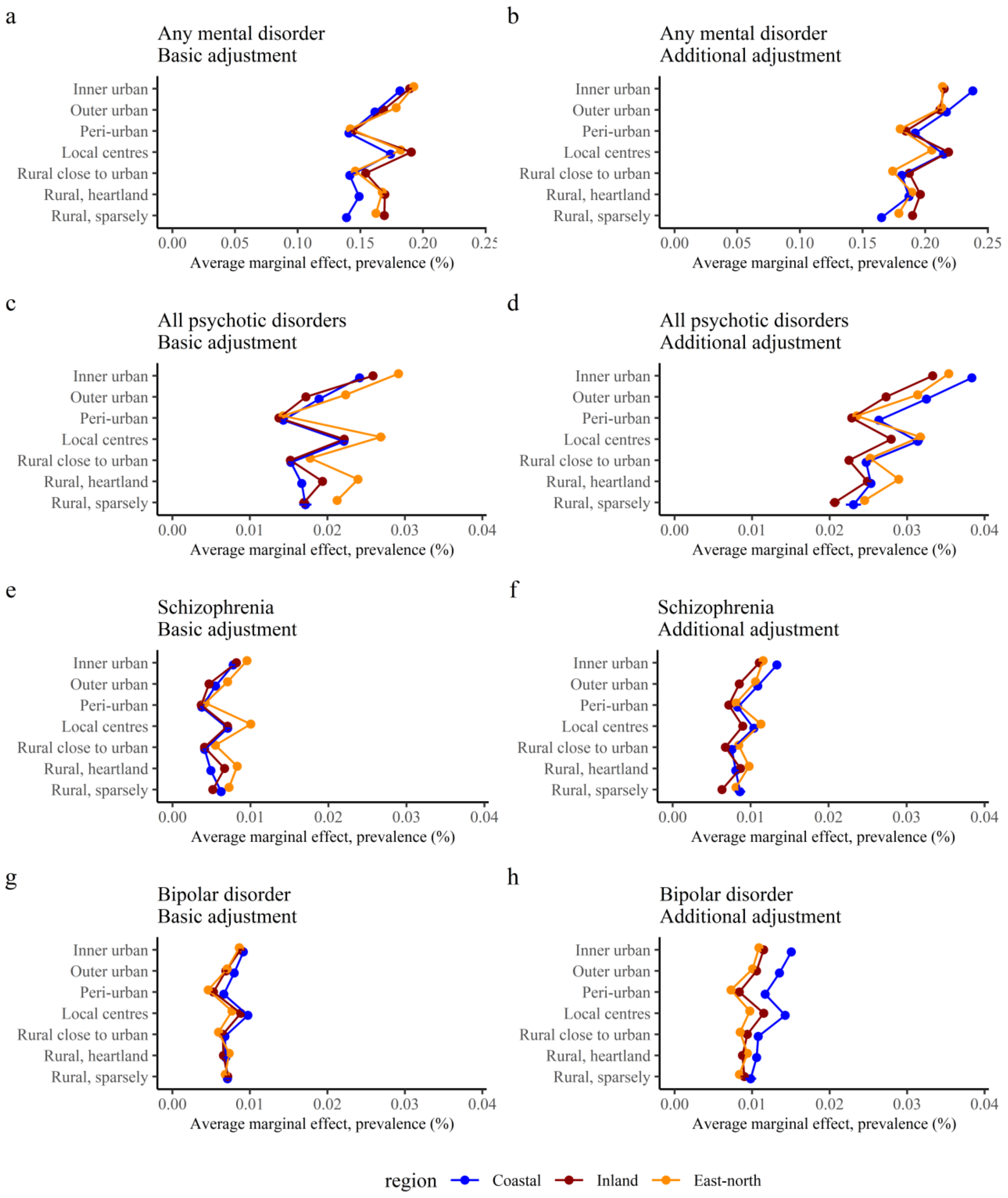

Coastal, Inland, and East-north regions are described in Figure 1d. In the basic adjustment, prevalence ratios are adjusted for age, gender, and calendar time. In the additional adjustment, prevalence ratios are adjusted for age, gender, calendar time, region, origin, residence history, household income, economic activity, and Charlson comorbidity index. Error bars indicate 95% CIs.

**Supplementary Figure S5: Prevalence ratios of selected mental disorders by place of residence in all treatment facilities and inpatient care only**

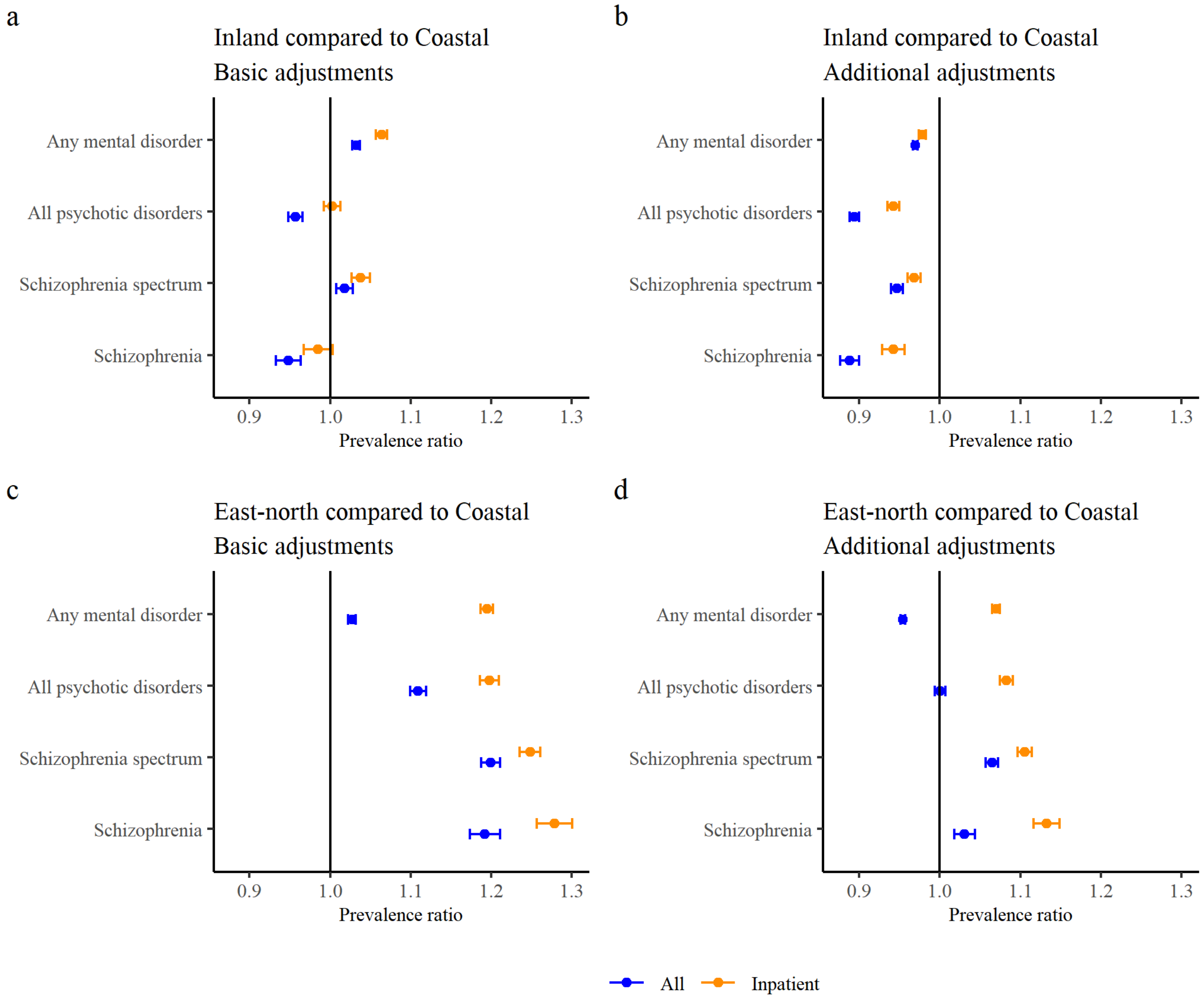

Coastal, Inland, and East-north regions are described in Figure 1d. In the basic adjustment, prevalence ratios are adjusted for age, gender, and calendar time. In the additional adjustment, prevalence ratios are adjusted for age, gender, calendar time, region, origin, residence history, household income, economic activity, and Charlson comorbidity index. Error bars indicate 95% CIs.

**Supplementary Figure S6: Prevalence ratios of selected mental disorders by place of residence and place of birth**

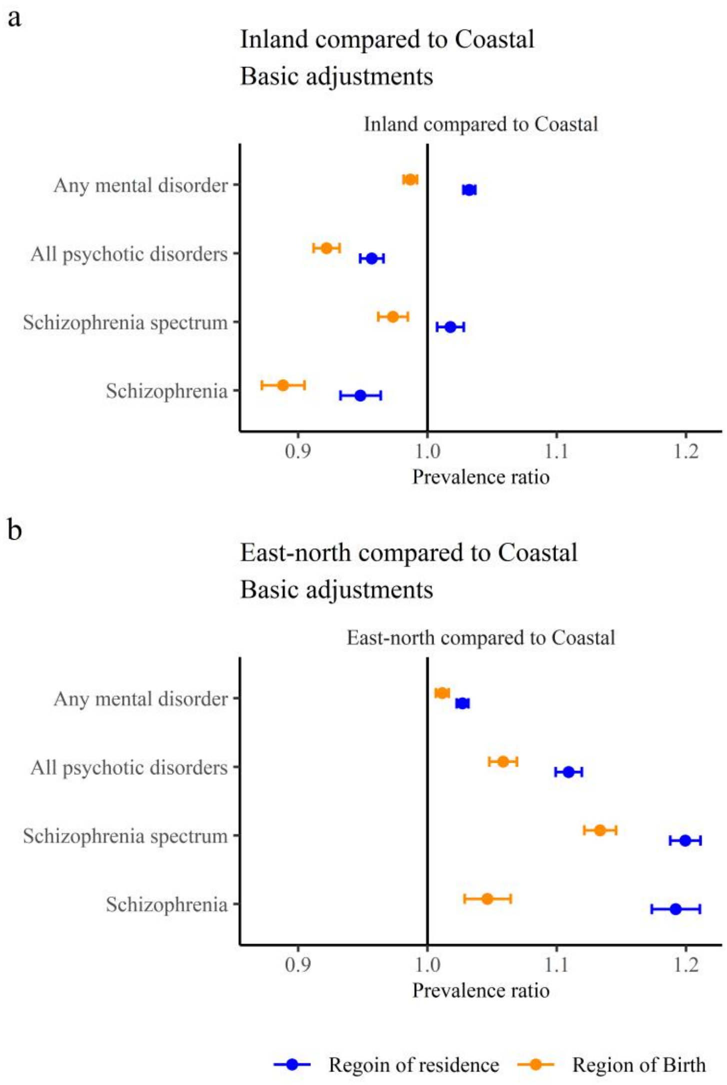

Coastal, Inland, and East-north regions are described in Figure 1d. In the basic adjustment, prevalence ratios are adjusted for age, gender, and calendar time. Error bars indicate 95% CIs.

**Supplementary Figure S7: Prevalence ratios and incidence rate ratios of inpatient treated mental disorders by place of residence**

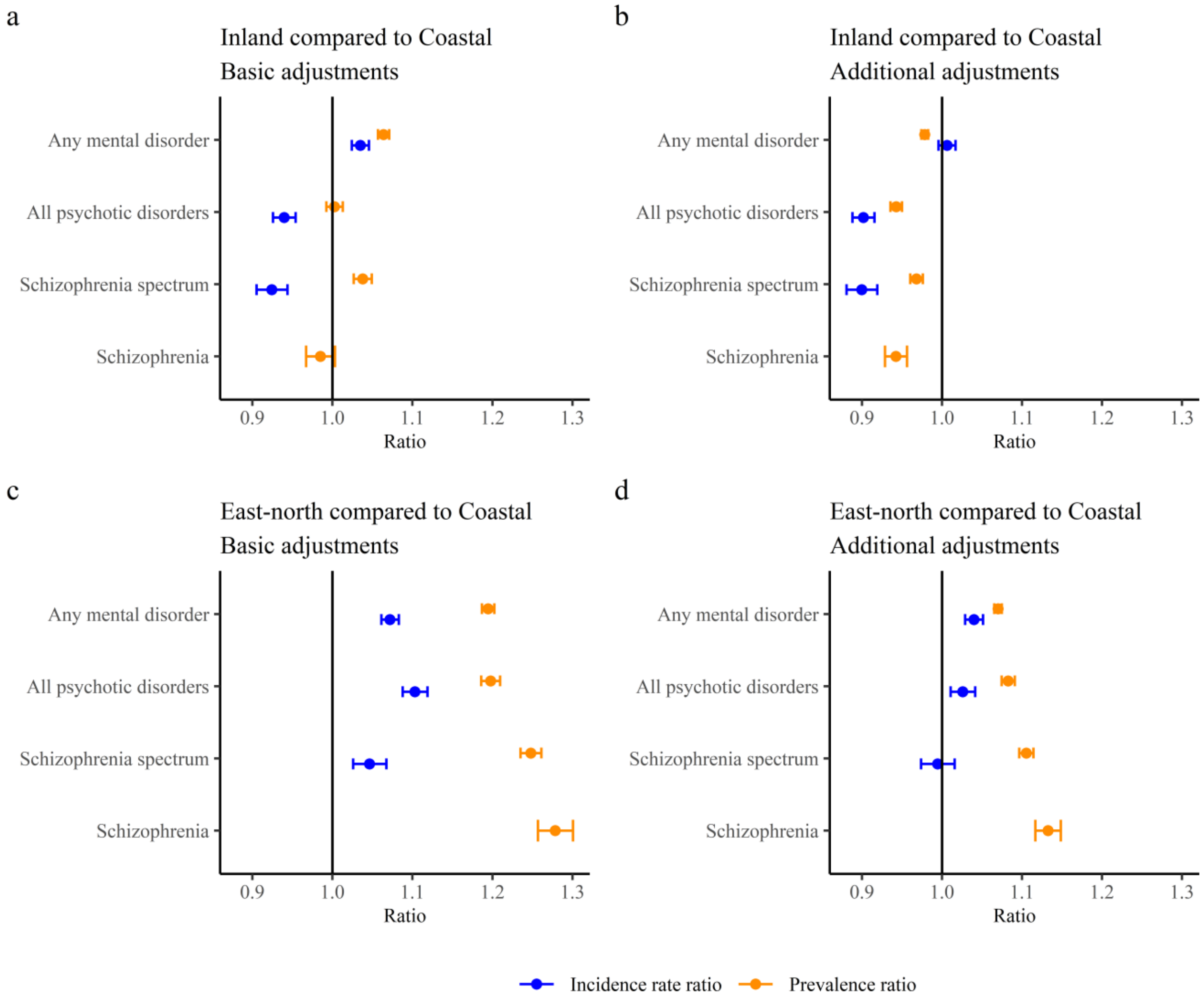

Coastal, Inland, and East-north regions are described in Figure 1d.In the basic adjustment, prevalence ratios are adjusted for age, gender, and calendar time. In the additional adjustment, prevalence ratios are adjusted for age, gender, calendar time, region, origin, residence history, household income, economic activity, and Charlson comorbidity index. Error bars indicate 95% CIs.

**Supplementary Figure S8: Prevalence ratios of selected mental disorders by place of residence, men and women analyzed separately**

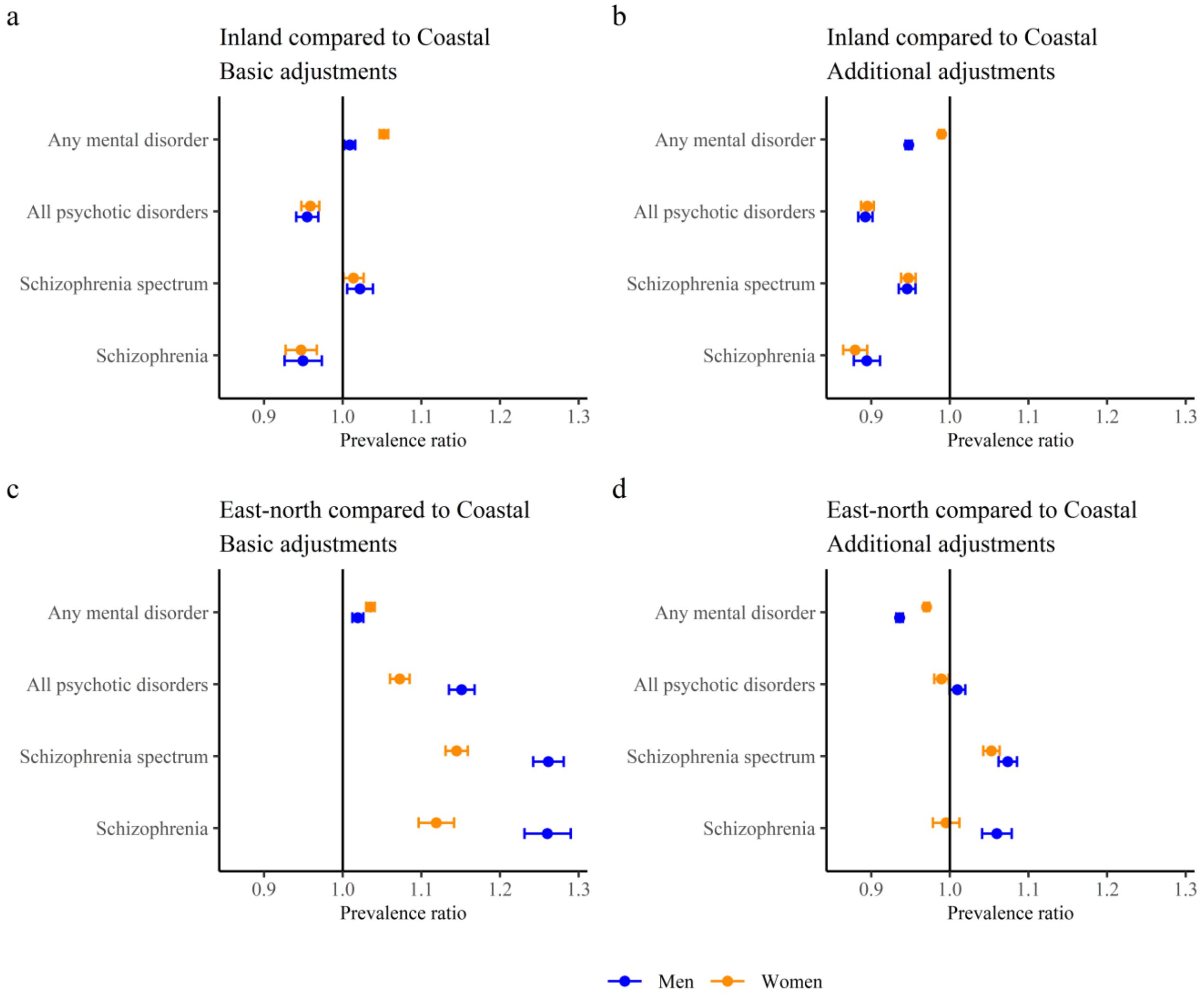

Coastal, Inland, and East-north regions are described in Figure 1d.In the basic adjustment, prevalence ratios are adjusted for age, gender, and calendar time. In the additional adjustment, prevalence ratios are adjusted for age, gender, calendar time, region, origin, residence history, household income, economic activity, and Charlson comorbidity index. Error bars indicate 95% CIs.
